# Experience of contraceptive care by midwives for nonpostpartum individuals in the Netherlands: a mixed methods study

**DOI:** 10.1101/2024.08.01.24311261

**Authors:** Merel Sprenger, Megan D. Newton, Renee N.N. Finkenflügel, Matty R. Crone, Jessica C. Kiefte-de Jong, M. Nienke Slagboom

**Author notes:** Corresponding author Merel Sprenger, Wijnhavengebouw, Turfmarkt 99, 2511 DP Den Haag, the Netherlands.

## Abstract

Since 2015, Dutch midwives have been authorised to prescribe all contraception. Initially providing contraceptive care to postpartum clients, they are increasingly offering it to anyone. It remains unknown how this broader population experiences this care. Therefore, this mixed methods study aims to explore experiences of nonpostpartum individuals receiving contraceptive care from Dutch primary care midwives.

At 13 midwifery practices in the Netherlands, participants were recruited to fill out a survey and participate in an in-depth semi-structured interview, both based on Levesque’s Conceptual Framework of Access to Health. Univariate and multivariate logistic regression analyses were applied to survey data (n = 91) and thematic analysis to interview data (n = 10).

Most survey participants (87.8%) received an intrauterine device during their appointment. A majority (58.2%) rated their care a 10 out of 10. Giving full marks was significantly associated with a higher perceived income (adjusted OR = 3.19, 95% CI = 1.21-8.81, p = 0.021), adjusted for appointment type and time since appointment. Participants reported receiving understandable information, being taken seriously, and having enough time during their appointment. Interviews revealed that participants especially appreciate how midwives make them feel at ease, midwives’ expertise, and the convenience of access.

To conclude, given the positive experiences reported by nonpostpartum individuals with contraceptive care from midwives, efforts should be made to improve task sharing and to increase awareness of midwives as contraception providers. Future research should compare care experiences across all types of providers and include a more representative population.

**Statement of significance:** 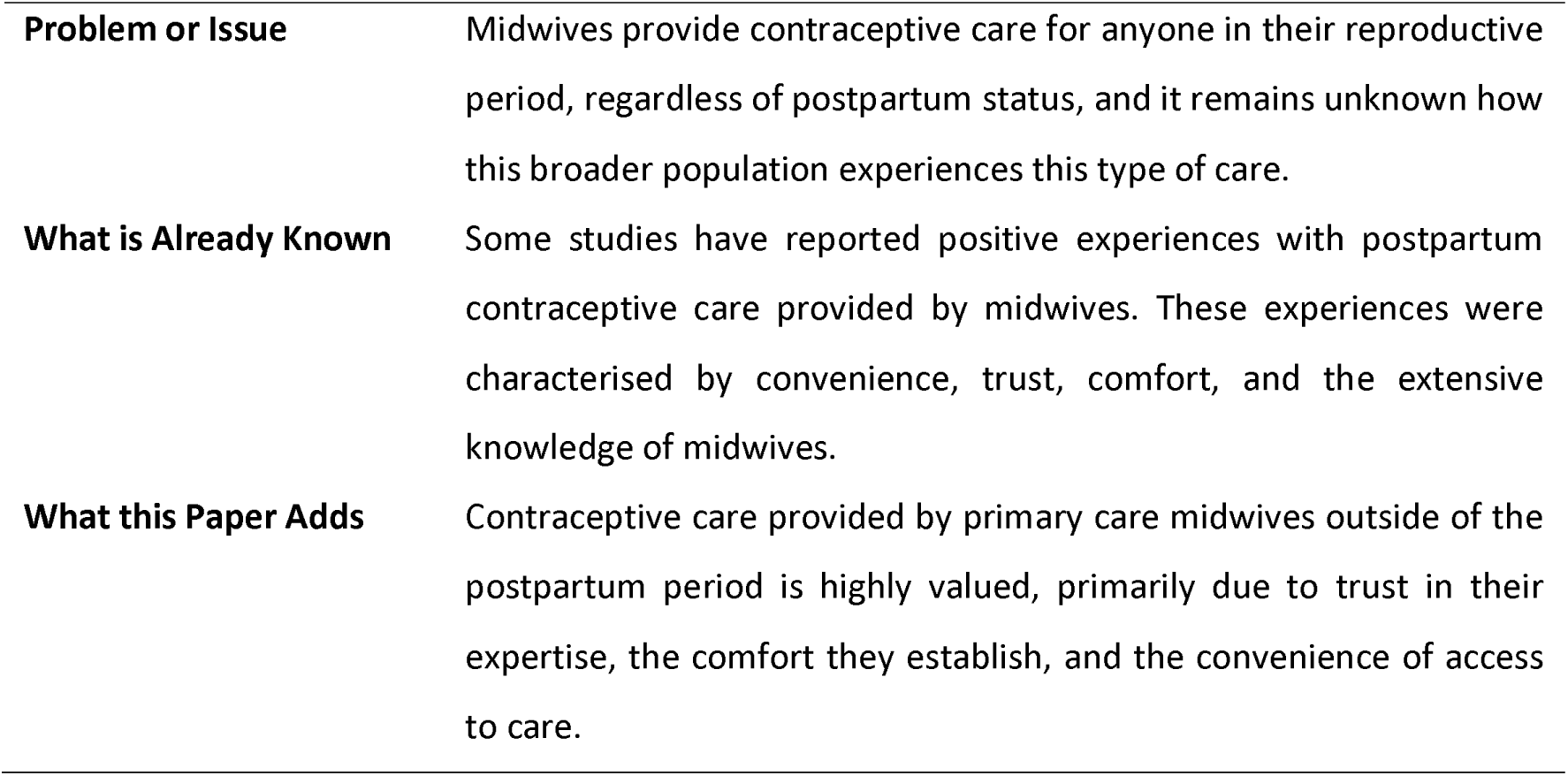

## Introduction

Contraception helps people in their reproductive years to exercise their right to decide if and when to have children (United Nations, 1995). Although widely available, there is not one perfect contraceptive method, not in terms of effectivity nor in terms of risks (Teal and Edelman, 2021). Moreover, while many different options exist, not everyone has the means, values or responsibilities to choose freely (Ross and Solinger, 2017). Therefore, access to appropriate, high quality contraceptive care and counselling plays an important role in the navigation of the biological and social reality of reproduction (Cadena et al., 2022).

In the Netherlands, people most often visit their general practitioner (GP) or GP assistant for contraceptive care, with GPs being the first contraceptive care provider for 82% of Dutch under 25-year-olds (van Ditzhuijzen et al., 2021). Additionally, contraceptive care is provided by doctors and nurses at abortion or sexual health clinics, and gynaecologists (Rutgers, 2024). In 2008, another healthcare professional was added to this list: midwives were authorised to insert intrauterine devices (IUDs). Since 2015, they also have the jurisdiction to prescribe all contraception methods including birth control pills (KNOV, 2022; Wagemakers and Vaandrager, 2015). Midwives have since primarily provided postpartum contraceptive care. Nevertheless, an increasing number of Dutch midwifery practices are expanding their services, offering contraceptive care to anyone in their reproductive period. This is in line with World Health Organization recommendations for family planning to improve access to contraceptive care through task sharing across professional cadres (WHO, 2017).

Internationally, healthcare systems are often not organised for midwives to be contraceptive providers. In Europe, Sweden is the only country where midwives play a primary role in family planning services, even providing 80% of all contraceptive counselling. Midwives are allowed to prescribe contraception in three other European countries: Estonia, the United Kingdom, and the Netherlands (Kopp Kallner et al., 2015; Sedlecky et al., 2020). Meanwhile, in the United States, midwives in Washington state recently (2022) gained prescriptive authority to prescribe contraception (Zell et al., 2024).

Some studies have reported positive experiences with postpartum contraceptive care provided by midwives. These experiences were characterised by convenience, trust, comfort, and the extensive knowledge of midwives (Carr et al., 2018; Walker et al., 2021). However, there have been hardly any studies on experience of contraceptive care by midwives for anyone in their reproductive period, regardless of postpartum status, and it remains unknown how this broader population experiences this type of care. The few studies that do not focus on the postpartum population explore the perspectives of midwives rather than the people receiving care (Höglund and Larsson, 2019; Kettyle and Klima, 2002; Kolak et al., 2017), or involve specific populations, like people with a migration background in Sweden (Kolak et al., 2022). In that study, trust was the main driver for accessing contraceptive counselling at the midwife. Additionally, lack of knowledge about contraception, the reproductive system, and the position of midwives as primary contraceptive providers in Sweden impacted their access to and experience of contraceptive care (Kolak et al., 2022).

### Objectives

The aim of this study is to explore experiences of contraceptive care at the primary care midwife for nonpostpartum individuals. More specifically, we want to know 1) how nonpostpartum individuals evaluate and experience contraceptive care at the primary care midwife; and 2) which sociodemographic factors and appointment characteristics are associated with this evaluation.

## Methods

### Study design

This is an explanatory mixed methods study, allowing for a comprehensive answer to our research questions through both quantitative and qualitative methods (Creswell, 2015). The relationship and sequence of the research components is presented in Figure 1.

**Figure 1.**
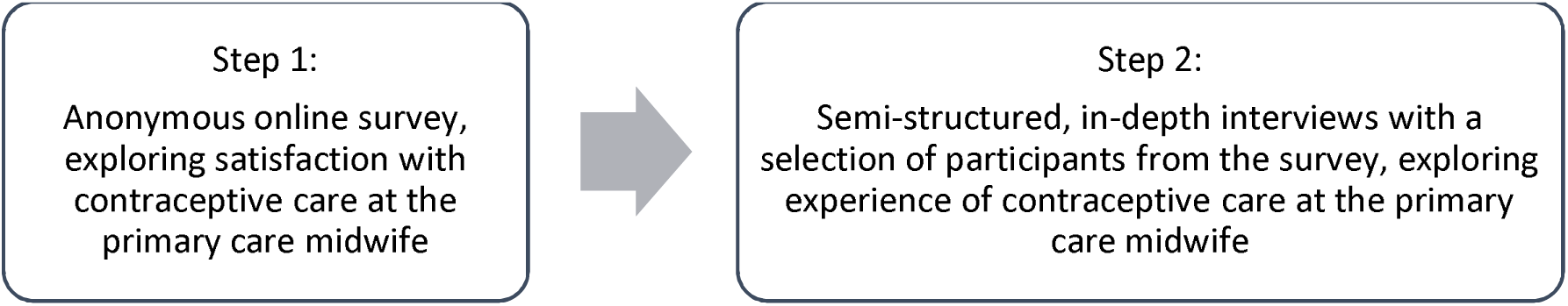
Diagram illustrating sequence of mixed methods research components.

The survey and interview questions were designed by MS in collaboration with RNNF, MRC, JCKJ, and MNS. To make sure we used suitable language, four midwives and two experts on contraception provided feedback on the survey. The survey was pilot tested by two medical students, including MDN, and by a PhD candidate on reproductive justice. Survey and interview questions are based on previous research on experience of contraceptive care at Dutch GPs and are grounded in Levesque’s Conceptual Framework of Access to Health (Levesque et al., 2013; van Ditzhuijzen et al., 2021). As presented in Figure 2, this framework consists of five dimensions of accessibility: 1) Approachability; 2) Acceptability; 3) Availability and accommodation; 4) Affordability; 5) Appropriateness. These dimensions provide information on the healthcare system, e.g., about available information, values, organisation, costs, and provider characteristics. Additionally, there are five abilities of individuals accessing healthcare, each related to one dimension: 1) Ability to perceive; 2) Ability to seek; 3) Ability to reach; 4) Ability to pay; and 5) Ability to engage. This framework has been used to explore experiences of by considering different dimensions of healthcare access and challenges to not only look at the characteristics of the healthcare system and the care provided, but to also recognise individual abilities that may influence access to care (Cu et al., 2021; Levesque et al., 2013).

**Figure 2.**
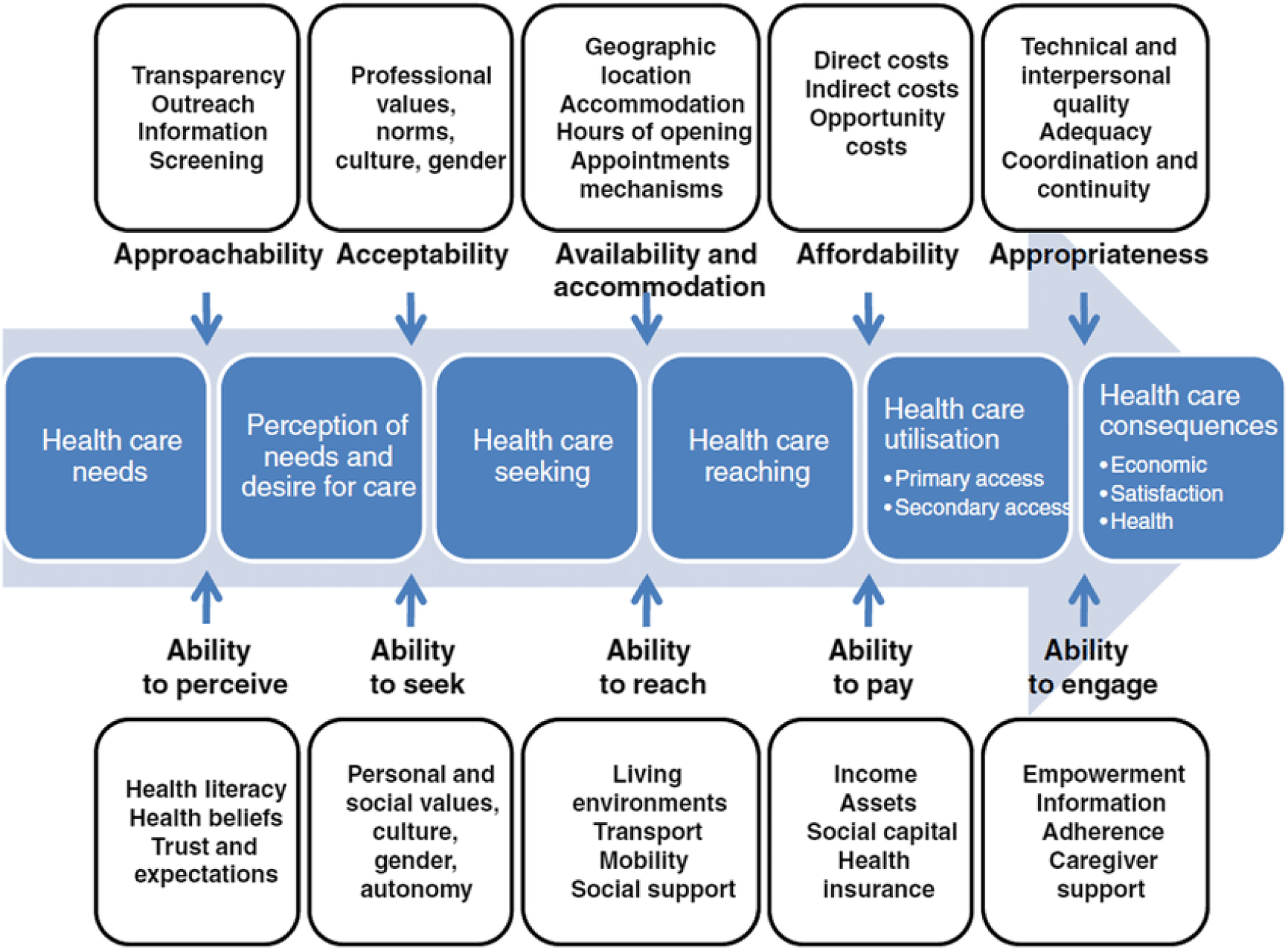
Levesque’s Conceptual Framework of Access to Health. (Levesque et al., 2013)

### Setting and data collection

From March 2024 to July 2024, participants were recruited through non-probability sampling at 13 midwifery practices and two ultrasound centres, providing contraception in both urban (n = 11) and rural (n = 4) areas of the Netherlands. Midwives informed people receiving contraceptive care through a flyer at the end of appointments or via social media. MN also visited practices to personally invite potential participants. People were eligible to take part if they were 16 years or older, accessed contraceptive care at the midwife, and had not given birth in the past six months. They also needed to understand Dutch or English, as the surveys were provided in these languages.

The flyer and social media post led to a webpage with additional information where participants could navigate to the consent section, followed by the anonymous survey. After completing the anonymous survey, participants could receive a voucher of €10 by contacting the research team. We then sent an interview information letter and asked if they would like to participate in an interview. We contacted participants who agreed to schedule an interview in person or via an online videocall, resulting in a convenience sample. After explaining the study and addressing questions, participants provided informed consent before the interview, including consent to audio recording. They could pause or stop the interview at any time and skip any question. Participants received a voucher of €15 after the interview.

### Units of study

The survey included questions on sociodemographic characteristics, previous experiences with contraceptive care, conversations about contraception, appointment characteristics, expectations and experiences, information expected and received, satisfaction, and costs. In the last section, participants rated their appointment from 0 (very bad) to 10 (very good) separately for advice and insertion/prescription. They also answered open-ended questions on what they appreciated and what could be improved. All survey questions were mandatory, but participants could choose ‘do not know’ or ‘rather not say’ if they wished to skip a question. The survey data that support the findings of this study are available from the corresponding author, MS, upon reasonable request. The survey can be found in Appendix A.

Ten people participated in a semi-structured in-depth interview; participants were not previously known to the researchers. One interview was conducted by MS and nine by MDN, both women. After interviews, field notes were made and discussed, including whether saturation had been reached. The interview consisted of two parts: first, mapping participants use of contraceptive methods across the life course, and second, addressing participants’ experience of contraceptive care at the midwife. We started by asking how participants accessed information about contraceptive care at the midwife, why they chose to go there and what they expected from the appointment. Next, participants walked us through their experience from the waiting room to leaving the midwifery practice, were prompted for more details if needed, and elaborated on what they appreciated and what could be improved. Finally, if applicable, participants were asked to compare their recent experience of contraceptive care at the midwife with earlier experiences with other providers (e.g., GP or gynaecologist). The interviews lasted an average of 50 minutes, ranging from 38 minutes to 1 hour and 12 minutes. Due to the nature of the research, interview data is not available. The entire interview guide can be found in Appendix B.

### Analysis

Quantitative analysis was performed using RStudio (version 2022.02.3+492) by MS. The answers “do not know” and “prefer not to answer” were coded as missing. As there was very little missing data, all participants were included in analyses and there was no need for multiple imputation (Jakobsen et al., 2017). Continuous data were presented with a mean and standard deviation (SD) or median and interquartile range (IQR), categorical data were presented as counts and percentages. To explore how nonpostpartum individuals evaluate contraceptive care at the primary care midwife, the outcome variable was composed of the grade for advice, the grade for insertion/prescription, or the mean of the two in case participants indicated they had received both advice and insertion/prescription. Based on the extremely negatively skewed distribution of this grade and majority of participants grading their appointment a 10, we decided to create a binary outcome variable, distinguishing between participants who gave full marks and those that did not. Next, to assess which sociodemographic factors and appointment characteristics are associated with this evaluation, we first performed univariate logistic regression analyses with sociodemographic variables and appointment characteristics. Then, variables were selected for multivariate logistic regression analysis if p-values were <.10 in the univariate logistic regression analyses. Log odds from regression analyses were transformed into odds ratios (ORs) with their respective 95% confidence interval (CI) to facilitate interpretability.

Interview recordings were transcribed verbatim and analysed using Atlas.ti. In our iterative coding process, we performed an integrated thematic analysis using both deductive coding based on Levesque’s Conceptual Framework of Access to Health and inductive coding, allowing for a more in-depth exploration of participants’ experiences. Coding was done by MDN, supervised by MS. This analysis of interviews helps explore the experience of contraceptive care of nonpostpartum individuals at the primary care midwife and explain the survey results.

### Ethical approval

The study was reviewed by the ethical committee and received a waiver from the Medical Research Ethics Committee of Leiden Den Haag Delft on 3 November 2021 under reference number N21.127 as it was not deemed to be subject to the Medical Research Involved Human Subjects Act.

## Results

In this mixed methods study on experiences of contraceptive care for nonpostpartum individuals at the primary care midwife, a description of participants is presented first, followed by our findings on evaluation and experience of care. Throughout, results will be linked to Levesque’s Conceptual Framework of Access to Health (Figure 2) (Levesque et al., 2013).

### Participants

Of the 162 people who opened the survey, 52 did not start filling it out, 18 started but did not complete it, and 92 participants completed the survey. 1 participant was excluded because they had given birth in the past 6 months, resulting in a sample of 91 survey participants. There was 1.4% of missing data across all variables. For the variable time since appointment, there were 8 participants with missing data. One participant did not give a grade for advice and one participant did not give a grade for insertion. Since these participants’ appointments included both advice and insertion, the grade they did provide was used as outcome data.

Participants had a mean age of 29 (SD: 6.87) in the survey and 28 (SD: 6.96) in the interviews (Table 1). A majority had a theoretical education, were born in the Netherlands, were not religious, and were in a relationship. Most survey participants reported a comfortable income. 69.2% of survey participants had ever visited a midwife and about a third had sought contraceptive care from a midwife before. Four interview participants had never been pregnant before. Of the six participants that had previously received prenatal care from a midwife, three visited the same midwife for contraception. All interview participants had either a hormonal intrauterine system (IUS, n = 6) or IUD (n = 4) inserted.

**Table 1.** Characteristics of participants for the survey and interviews.

| Participant characteristic | Survey (n = 91) | Interviews (n = 10) |
| --- | --- | --- |
|  | n (%) | n (%) |
| Age, mean (SD) | 29.00 (6.87) | 27.80 (6.96) |
| Education |  |  |
| Practical | 26 (28.9) | 3 (30.0) |
| Theoretical | 64 (71.1) | 7 (70.0) |
| Themselves and parents born in the Netherlands | 82 (90.1) | 8 (80.0) |
| Religion |  |  |
| No religion | 74 (82.2) | 8 (80.0) |
| Christian | 16 (17.8) | 2 (20.0) |
| Income |  |  |
| Difficult or coping | 32 (36.4) |  |
| Comfortable | 56 (63.6) |  |
| In a relationship | 82 (90.1) | 10 (100.0) |
| Has children |  | 6 (60.0) |
| Ever visited a midwife | 63 (69.2) | 10 (100.0) |
| Contraception providers visited before |  |  |
| Midwife | 33 (36.3) | 3 (30.0) |
| General practitioner | 77 (84.6) | 8 (80.0) |
| Gynaecologist | 18 (19.8) | 4 (40.0) |

### Evaluation of care and associated characteristics

Most survey participants (86.8%) had contraception prescribed or inserted and when they did, they often chose an IUS or IUD (87.8%), see Table 2. Their experience was incredibly positive, with a median of 10 on a 1-10 scale (range = 5-10) and with 58.2% of participants grading it a 10. In the interviews, one participant reflected on her positive score by saying: “Then I will just give a 10. I really can’t think of anything that did not go well or what could have been done differently.”(P8) This positive evaluation is further illustrated in answers to other questions about experience within the Appropriateness dimension and the Ability to engage, with 89.0% finding the information during the appointment very understandable, 95.6% feeling at ease, 96.7% feeling like they were taken seriously, and 98.9% feeling like there was enough time for the appointment.

**Table 2.** Appointment characteristics and experience from survey data (n = 91).

| **Appointment characteristic** | **n (%)** |
| --- | --- |
| Type of appointment |  |
| Advice only | 12 (13.2) |
| Insertion/prescription only | 15 (16.5) |
| Advice and insertion/prescription | 64 (70.3) |
| Time since appointment |  |
| Less than 1 week ago | 30 (36.1) |
| Between 1 week and 2 months ago | 17 (20.5) |
| More than 2 months ago | 36 (43.4) |
| Contraception in month before appointment |  |
| No | 56 (64.4) |
| Yes | 31 (35.6) |
| Area of midwifery practice |  |
| Urban | 51 (56.0) |
| Rural | 40 (44.0) |
| Role of costs in choice of contraception provider |  |
| No role | 45 (50.6) |
| Small role | 34 (38.2) |
| Large role | 10 (11.2) |
| Method of contraception chosen* |  |
| Combined pill | 2 (2.2) |
| Progesterone-only pill | 1 (1.1) |
| Intrauterine system (IUS) (hormonal) | 54 (60.0) |
| Intrauterine device (IUD) (copper) | 25 (27.8) |
| Contraceptive implant | 5 (5.6) |
| Male condom | 3 (3.3) |
| Natural family planning | 1 (1.1) |
| Sterilisation | 2 (2.2) |
| Grade, median (IQR) | 10 (9-10) |
| Grade |  |
| <10 | 38 (41.8) |
| 10 | 53 (58.2) |

\*Participants could choose up to two methods of contraception.

With regards to the Ability to perceive, for all subjects asked about, participants received information more often than they had expected to. Notably, 59.3% received information contraception effectiveness in preventing pregnancy, while only 47.3% expected this. Information on side effects seemed to be more important and had the smallest difference between expectation and experience, with 75.8% expecting information and 79.1% having received information. The largest difference was for costs, with 34.1% expecting information and 52.7% having received information on costs. Related to Acceptability, most received care from female midwives (97.8%)and while some participants had no preference for their contraception provider’s gender, most preferred a female provider both for advice (68.1%) and insertion (83.5%). An interview participant elaborated on this, referring to women’s embodied knowledge: “Women amongst themselves, you truly understand what you are talking about. Whether it’s about inserting contraception or getting contraception, in whatever form. I think that, by and large, men have less experience with that.”(P3)

In the univariate logistic regression analyses, perceived income, appointment type, and time since appointment had a p-value of <0.10 and hence were included in the multivariate logistic regression analysis (Table 3). In this model, only the association between perceived income and giving full marks remained significant, with an adjusted odds ratio of 3.19 (95% CI (1.21-8.81), p = 0.021). This means that, after adjusting for time since appointment and appointment type, people with a comfortable perceived income have more than three times the odds of reporting full marks compared to people who report to have a difficult perceived income or are coping. This finding may therefore be related to the Ability to pay.

**Table 3.** Univariate and multivariate logistic regression analyses for sociodemographic and appointment determinants of giving full marks for contraceptive care experience at the primary care midwife.

| Determinant | Univariate |  | Multivariate (n = 79) |  |
| --- | --- | --- | --- | --- |
|  | OR (95%CI) | P-value | aOR (95%CI) | P-value |
| Age | 0.99 (0.93-1.05) | 0.793 |  |  |
| Education |  |  |  |  |
| Practical | Ref. |  |  |  |
| Theoretical | 1.07 (0.42-2.69) | 0.883 |  |  |
| Ethnicity |  |  |  |  |
| (Parents) born in the Netherlands | Ref. |  |  |  |
| (Parents) born abroad | 0.54 (0.13-2.18) | 0.382 |  |  |
| Religion |  |  |  |  |
| Not religious | Ref. |  |  |  |
| Christian | 0.50 (0.16-1.49) | 0.215 |  |  |
| In a relationship |  |  |  |  |
| No | Ref. |  |  |  |
| Yes | 0.37 (0.05-1.62) | 0.226 |  |  |
| Perceived income |  |  |  |  |
| Difficult or coping | Ref. |  | Ref. |  |
| Comfortable | 3.09 (1.27-7.76) | 0.014 | 3.19 (1.21-8.81) | 0.021 |
| Midwife area |  |  |  |  |
| Urban | Ref. |  |  |  |
| Rural | 0.95 (0.41-2.20) | 0.899 |  |  |
| Ever visited a midwife |  |  |  |  |
| No | Ref. |  |  |  |
| Yes | 1.07 (0.43-2.62) | 0.887 |  |  |
| Time since appointment |  |  |  |  |
| Less than two months | Ref. |  | Ref. |  |
| More than two months | 0.41 (0.17-1.00) | 0.052 | 0.43 (0.16-1.11) | 0.084 |
| Contraception in month before appointment |  |  |  |  |
| No | Ref. |  |  |  |
| Yes | 1.47 (0.60-3.70) | 0.407 |  |  |
| Appointment type |  |  |  |  |
| Insertion and advice or insertion only | Ref. |  | Ref. |  |
| Advice only | 0.31 (0.08-1.06) | 0.071 | 0.35 (0.08-1.31) | 0.129 |
| Previous contraception at midwife |  |  |  |  |
| No | Ref. |  |  |  |
| Yes | 1.17 (0.49-2.82) | 0.730 |  |  |
| Previous contraception at GP |  |  |  |  |
| No | Ref. |  |  |  |
| Yes | 0.74 (0.21-2.35) | 0.619 |  |  |
| Previous contraception at gynaecologist |  |  |  |  |
| No | Ref. |  |  |  |
| Yes | 1.16 (0.41-3.47) | 0.783 |  |  |

### Experience of care

89 participants responded to the open survey question elaborating on what they appreciated and commonly responded that the midwife scheduled plenty of time for the appointment, made participants feel at ease, clearly explained every step of IUD insertion, and was familiar because of previously received care. As for what participants found unpleasant, 21 wrote nothing or responded with a hyphen or x, 47 participants explicitly noted that nothing was unpleasant, while 12 noted that the IUD insertion hurt.

To further understand the experience of contraceptive care at the midwife, we now present the results of the ten in-depth interviews with nonpostpartum individuals who had visited a midwife for IUD insertion. In the analyses of the interviews, four themes emerged: feeling at ease, convenience of access, trust in midwife as expert, and preconception of pregnancy being midwives’ preoccupation. An overview of the findings in light of the dimensions and corresponding abilities of Levesque’s Conceptual Framework of Access to Health (Figure 2) is presented in Table 4 (Levesque et al., 2013).

**Table 4.** Overview of findings in light of Levesque’s Conceptual Framework of Access to Health.

| Dimension, ability | Approachability, Ability to perceive | Acceptability, Ability to seek | Availability and accommodation, Ability to reach | Affordability, Ability to pay | Appropriateness, Ability to engage |
| --- | --- | --- | --- | --- | --- |
| Survey |  | - 83.5% prefer a female provider for insertion<br>- Midwife was familiar because of previous visits | - 98.9% felt there was enough time for the appointment | - People with a more comfortable perceived income have more than three times the odds of reporting full marks, adjusted for time since appointment and appointment type | - 89.0% found information during appointment very understandable<br>- 95.6% felt at ease<br>- 96.7% felt taken seriously<br>- More information received than expected<br>- Midwife clearly explained every step |
| Interviews | *Trust in midwife as expert*<br>- Midwife is familiar with female body<br>- Midwife often inserts IUDs<br>*Presumption of pregnancy being midwives' preoccupation*<br>- Not widely known that midwives provide contraception | *Presumption of pregnancy being midwives' preoccupation*<br>- Common idea that midwife is for pregnancy care only | *Feeling at ease*<br>- Midwife took enough time<br>- Midwife could perform an ultrasound<br>*Convenience of access*<br>- Short travel distance<br>- Appointment was booked within reasonable time | *Convenience of access*<br>- Lower costs than gynaecologist<br>- Indirect costs: short travel distance | *Feeling at ease*<br>- Midwife clearly explained each step of the procedure and asked consent before proceeding<br>- Midwife made participants feel like a person<br>- At ease because of expertise and frequency of doing this<br>- Could raise concerns, were listened to well<br>- Open, friendly, personal |

#### Feeling at ease

The theme ‘feeling at ease’ reflects the overarching positive experience of the participants with regards to the dimensions of Appropriateness, Ability to engage, and Availability and accommodation. Participants attributed feeling at ease to the midwives’ approach, which was characterised by managing expectations and consent, taking a personal approach, taking time, and performing an ultrasound after IUD insertion.

##### Managing expectations and consent

Not all participants had expected the midwife to fully explain the procedure before the insertion. Nevertheless, an appreciation of knowing what to expect was a reoccurring theme, as it allowed them to understand what was going to happen and to get comfortable with the midwife before the insertion. Moreover, before each next step during the insertion procedure, the midwife would explain what was going to happen and ask for consent before proceeding. This was appreciated because the participants knew what was happening and it made them feel involved in the procedure, like they were doing it together. They were also told in advance when something might be painful. This was seen as a positive thing because they knew what to expect. One participant, who reported a very positive experience of IUD insertion, elaborated that the explanation and consent process played a significant role in making her feel at ease, she said that “each step of the way, she would genuinely check in with me every time if I was okay, and ask if she could continue when she saw that it was unpleasant.”(P9)

##### Personal approach

Three participants expressed feeling that the midwife saw them as a person rather than a number, especially compared to earlier experiences at the gynaecologist: “In the hospital they don’t know, there you are just the umpteenth patient number on a day. They don’t know what happened in terms of children, childbirth and the like.”(P6) Compared to their previous IUD insertion at the GP, two participants shared they did not feel seen as a person and felt rushed. The midwife really made them feel valued as an individual. Having a prior relationship helped, as is illustrated by the following quote: “I don’t actually know my GP that well, I barely ever visit. And this midwife, I knew very well, so I think I also had a better relationship than with my own GP.”(P5) Participants further noted the midwife’s openness, friendliness, and personal approach, sharing her own experiences, which put them at ease. One participant shared that the IUD insertion became a more positive experience because the midwife seemed to enjoy her job, which was different compared to the GP where she had felt more like a burden.

##### Time

The feeling of enough time that the participants experienced mostly stemmed from not feeling rushed by the midwife. They felt like the midwife took the time to explain all the information and to answer all their questions. Especially when reflecting on the invasive and vulnerable procedure of an IUD insertion, one participant phrased this as follows: “It is not like: you walk in, it (the IUD, red.) is pushed in and you’re out the door again, so to speak. They really take their time with you.”(P10) All the participants stated that they felt like they could raise concerns if they had them and felt like they were listened to well. Compared to for example the GP, the participants reported that they had more time at the midwife and felt less rushed.

##### Ultrasound

Something that was also widely appreciated was the fact that the midwives performed an ultrasound either directly after insertion, or six weeks after the insertion. This gave participants a feeling of reassurance that their IUD/IUS was correctly inserted. They compared this to the GP where an ultrasound is not done and preferred to have this additional examination: “She checked with the ultrasound if it was properly in place. So that was also nice to immediately get that confirmation.”(P8)

#### Convenience of access

The theme ‘convenience of access’ explores the logistics around Availability and accommodation, Ability to reach, and Affordability that participants encountered when visiting a midwife as a contraception provider. The participants appreciated the fact that they did not have to go into hospital and that they did not have to travel far: “The midwife is nice and close to home, more accessible to call. Yes, then a hospital is further away.”(P6) In addition, participants felt like they were able to book an appointment at the midwife within a reasonable time. One participant also noted that they were able to book an appointment in the evening and that this would not have been possible at the GP. Finally, participants deemed the costs of seeking contraceptive care from a midwife lower than the gynaecologist, while comparable to the GP.

#### Trust in midwife as expert

The theme ‘trust in midwife as expert’ reflects the Approachability of and Ability to perceive midwives as contraceptive providers. Participants trusted the midwife’s expertise, both in terms of knowledge and skills, often linking this trust to the midwife’s familiarity with female anatomy and frequency of IUD insertions. One interviewee, for example said: “She is so experienced in placing those things (IUDs, red.), who knows how many times in a week! You gain so much more experience and are so much more skilled in it.”(P7) Participants frequently compared the midwife with the GP:

> “I feel like the midwife does insert contraception or at least IUDs more often than the GP. Of course, the GP has a much larger range of duties, and the midwife also has a broad range of duties, but that is mainly focused on pregnancy, childbirth, ‘uterus stuff.’ (…) And they just know a lot and I know that they can do it well, inserting contraception. So, I did have a much better or safer feeling compared to my GP.”(P5)

#### Presumption of pregnancy being midwives’ preoccupation

The themes above illustrate the participants’ explanations of their positive experiences at the midwife, but participants also reflected on potential disadvantages, related to the dimensions of Acceptability, Ability to seek and Approachability. The main disadvantage was the presumption that midwives only provide pregnancy care, not contraception. Consequently, although described by four participants, a certain meaning is ascribed when someone is seen at a midwifery practice. Two participants mentioned that around their appointment, one in the waiting room and one after, someone asked if they were pregnant:

> “And I received a text afterwards from someone who happened to see me there saying: hey, you were at the midwife, do you have to tell me something? And I said yes, I got an IUD, so there is nothing. That is kind of a small-town thing in our area. Like to just check, and that a midwife is immediately associated with you being pregnant. And that clearly doesn’t have to be the case.”(P3)

A third participant mentioned this as a hypothetical situation, and another had her own prejudices that only changed while on placement for her midwifery degree:

> “At first, when the GP suggested (to go to a midwife for IUD insertion, red.), I thought: I’m not going, the midwife is something for women who are slightly older or for women who just got pregnant and want contraception after that. Until I also saw younger women (come for contraception, red.) on placement. Then I thought: actually, this is also just a place to get it done as a younger woman.”(P4)

While these participants did not mind explaining that they were at the midwife for contraception, they mentioned not everyone might feel comfortable doing so. Although not described by all participants, these stories reiterate the idea that midwives are for pregnant people only. Hence, it is no surprise that all Dutch participants shared that many people are unaware of the possibility of contraceptive care at the midwife. On the other side, for the two participants who grew up outside of the Netherlands, the Dutch healthcare system was new and after searching for IUD care online and finding the midwife, they assumed this is how contraceptive care is organised in the Netherlands.

## Discussion

This mixed-methods study explored how nonpostpartum individuals evaluate and experience their contraceptive care at Dutch primary care midwives. The survey showed that participants evaluate this with high grades, with people with a more comfortable perceived income more often giving full marks. The interviews identified feeling at ease, convenience of access, and trust in the midwives’ expertise as common themes in participants’ explanations of their positive experiences during for IUD insertion appointments. These findings are comparable to other studies on contraceptive care at midwives where convenience (Carr et al., 2018; Walker et al., 2021), trust (Kolak et al., 2022; Walker et al., 2021), comfort, and midwives’ knowledge were appreciated(Walker et al., 2021). Although international studies on contraceptive care experiences at primary care providers, reproductive healthcare providers, and certified nurse midwives have also reported positive experiences, the ratings in this first study on contraceptive care from midwives in the Netherlands were remarkably high (Dehlendorf et al., 2017; Dehlendorf et al., 2016; Hart et al., 2023).

Table 4 presents an overview of our findings in light of Levesque’s Conceptual Framework of Access to Health. We will discuss these findings with regards to existing literature. As for the Acceptability dimension and the Ability to seek, some interview participants experienced prejudice about them being pregnant after being seen at a midwife. However, they did not mind explaining the actual reason for their visit, which is in contrast with findings from Kolak et al. where immigrant women who migrated to Sweden from outside of Europe feared their parents would find out they had visited a midwife for contraception (Kolak et al., 2022). Additionally, our survey and interview findings show that midwives are acceptable contraceptive care providers for two reasons in addition to their expertise. First, most participants preferred a female provider for contraceptive care, which is in line with the preference for female primary care physicians and gynaecologists (Spaich et al., 2019; Walker et al., 2024). Second, even though our participants were not postpartum at the time of their appointment, if they received care from a midwife before, they had already established a relationship, which is known to be of great value (Kolak et al., 2022; Walker et al., 2021). With regards to Availability and accommodation and the Ability to reach, both research methods showed midwives plan sufficient time for appointments, something that has also been associated with satisfaction of contraceptive care at Dutch GPs (van Ditzhuijzen et al., 2021). Furthermore, with regards to Affordability and the Ability to pay, we found that adjusted for appointment type and time since appointment, full marks were more often given by those with a higher perceived income. An association between income and quality of contraceptive care has previously been found in the United States, where affordability was also found to be a large barrier in accessing contraceptive care (Newton-Levinson et al., 2024; Wingo et al., 2023). Finally, our findings relating to the Ability to engage with and the Appropriateness of midwives as contraceptive providers Including time, trust, expertise, positive interactions, and a supportive environment have previously been highlighted as key factors in contraceptive care both at midwives and other providers (Fulcher et al., 2021; Kolak et al., 2022; Reed et al., 2022; van Ditzhuijzen et al., 2021).

While all five dimensions from the framework appeared in our study, the Appropriateness dimension emerged most often. This is not surprising, as according to the Levesque Conceptual Framework of Access to Health, this dimension and the Ability to engage become relevant in particular after having accessed care and these concepts consequently influence satisfaction, or experience with care (Levesque et al., 2013). Concurrently, these concepts, related to midwives’ interpersonal skills, being most relevant is in line with studies on another care experience when one might feel vulnerable: the mammogram. Interpersonal skills and positive attitude of mammography staff are important factors related to satisfaction of care, and explanation before and during the procedure resulted in fewer women experiencing pain during the mammogram (Almog et al., 2008; Van Goethem et al., 2003; Whelehan et al., 2016).

### Strengths

Through this study’s mixed methods design, we were able to both quantitatively study the evaluation of contraceptive care at the primary care midwife and perform a qualitative in-depth exploration of experience of care. Our questions were based on Levesque’s Conceptual Framework of Access to Health (Levesque et al., 2013), providing us with structure and a theoretical foundation, and on previous mixed methods work on contraceptive care at Dutch GPs (van Ditzhuijzen et al., 2021). The study fills a gap in the literature, as there are barely any studies internationally, and to our knowledge none in the Netherlands, on midwives as a contraceptive provider for all and more specifically not on people that did not receive this type of care as a continuation of prenatal care after delivery. A final strength is that we purposefully sampled our interview participants to ensure they had not received postpartum contraceptive care at the midwife.

### Limitations

This study has several limitations. First, related to recruitment, although we aimed to include only nonpostpartum individuals in both the survey and interviews, some midwife practices shared a social media post to inform clients about the study. It is likely that this was the reason a larger than expected proportion of survey participants reported their appointment to be more than two months ago, and why this variable had most missing data. Fitting our inclusion criteria and not being postpartum at the time of survey participation, it is possible that these participants could have been postpartum during their appointment. Secondly, our sample is relatively homogenous, especially when it comes to education, ethnicity, religion, and income. As a result, we have unfortunately not been able to explore in the interviews why people with a more comfortable perceived income more often evaluated their appointment with full marks. Our homogenous sample might be the result of our non-probability sampling and recruitment methods. Subsequently, this limits the generalisability of our survey findings. It is possible there was selection bias, resulting in more people with a positive experience taking part, both for the survey and interviews. Or these groups might not seek contraceptive care at midwives so often, as our results showed that the midwife is not well known as a contraception provider.

### Implications

Various implications follow from this study. First, given recommendations from the World Health Organization on access to contraception, it might be an option to implement task sharing and increasingly position midwives as contraceptive providers, specifically for IUD/IUS insertion and both in and outside of the Netherlands (WHO, 2017). To realise this, more midwives should be trained to provide contraception and awareness should be raised about midwives serving as primary care contraception providers. Second, since the experience of contraceptive care provided by midwives seems to be so positive, other providers could draw inspiration from the characteristics most appreciated, if they are not already implementing them: asking consent before each step, scheduling enough time, and taking a personal approach. Finally, a comparative study of the experience of contraceptive care at all available contraceptive providers should be performed, including a representative population.

## Conclusion

This study demonstrated that contraceptive care provided by primary care midwives in the Netherlands is highly valued, primarily due to trust in their expertise, the comfort they establish, and the convenience of access to care. Despite the presumption that midwives focus solely on pregnancy care, our findings reveal they are a suitable contraception provider for all. This presents an opportunity for task sharing of contraceptive care between midwives and other providers, particularly for IUD insertions.

## Data Availability

Survey data produced in the present study are available upon reasonable request to the authors. The survey can be found in Appendix A. Interview data is not available, but the interview guide can be found in Appendix B.

## Declaration of Generative AI and AI-assisted technologies in the writing process

Statement: During the preparation of this work the authors used ChatGPT in order to improve flow and conciseness of the content. After using this tool/service, the authors reviewed and edited the content as needed and take full responsibility for the content of the publication.

## Abbreviations

GP: General practitioner
IUD: Intrauterine device
IUS: Intrauterine system

## References

Almog, R., Hagoel, L., Tamir, A., Barnett, O., Rennert, G., 2008. Quality control in a national program for the early detection of breast cancer: women’s satisfaction with the mammography process. Women’s Health Issues 18:110–17. doi: 10.1016/j.whi.2007.12.007

Cadena, D.S., Chaudhri, A., Scott, C., 2022. Contraceptive care using reproductive justice principles: beyond access. American Journal of Public Health 112:S494–S99. doi: 10.2105/AJPH.2022.306915

Carr, S.L., Singh, R.H., Sussman, A.L., Rogers, R.G., Pereda, B., Espey, E., 2018. Women’s experiences with immediate postpartum intrauterine device insertion: a mixed-methods study. Contraception 97:219–26. doi: 10.1016/j.contraception.2017.10.008

Creswell, J.W., 2015. A concise introduction to mixed methods research. SAGE Publications, Incorporated, Thousand Oaks, California.

Cu, A., Meister, S., Lefebvre, B., Ridde, V., 2021. Assessing healthcare access using the Levesque’s conceptual framework– a scoping review. International Journal for Equity in Health 20:116. doi: 10.1186/s12939-021-01416-3

Dehlendorf, C., Anderson, N., Vittinghoff, E., Grumbach, K., Levy, K., Steinauer, J., 2017. Quality and content of patient–provider communication about contraception: differences by race/ethnicity and socioeconomic status. Women’s Health Issues 27:530–38. doi: 10.1016/j.whi.2017.04.005

Dehlendorf, C., Henderson, J.T., Vittinghoff, E., Grumbach, K., Levy, K., Schmittdiel, J., Lee, J., Schillinger, D., Steinauer, J., 2016. Association of the quality of interpersonal care during family planning counseling with contraceptive use. American Journal of Obstetrics and Gynecology 215:78.e1-78.e9. doi: 10.1016/j.ajog.2016.01.173

Fulcher, K., Archibald, A., Francoeur, J., 2021. ’They really hear you out’: lessons on providing contraceptive care from a community-based sexual health clinic. The Canadian Journal of Human Sexuality 30:243–51. doi: 10.3138/CJHS.2021-0018

Hart, L., Parsons, G., Beaudoin, J., Alonge, O., Karp, C., 2023. A mixed methods study of contraceptive counseling and care at a federally qualified health center in Maryland. Journal of Patient Experience 10. doi: 10.1177/23743735231183572

Höglund, B., Larsson, M., 2019. Midwives’ work and attitudes towards contraceptive counselling and contraception among women with intellectual disability: focus group interviews in Sweden. The European Journal of Contraception & Reproductive Health Care 24:39–44. doi: 10.1080/13625187.2018.1555640

Jakobsen, J.C., Gluud, C., Wetterslev, J., Winkel, P., 2017. When and how should multiple imputation be used for handling missing data in randomised clinical trials – a practical guide with flowcharts. BMC Medical Research Methodology 17:162. doi: 10.1186/s12874-017-0442-1

Kettyle, E.P., Klima, C., 2002. Adolescent emergency contraception: attitudes and practices of certified nurse-midwives. Journal of Midwifery & Women’s Health 47:68–73. doi: 10.1016/s1526-9523(02)00219-2

KNOV, 2022. Algemene maatregel van bestuur. Available from: https://www.knov.nl/werk-en-organisatie/informatie/algemene-maatregel-van-bestuur

Kolak, M., Jensen, C., Johansson, M., 2017. Midwives’ experiences of providing contraception counselling to immigrant women. Sexual & Reproductive Healthcare 12:100–06. doi: 10.1016/j.srhc.2017.04.002

Kolak, M., Löfgren, C., Hansson, S.R., Rubertsson, C., Agardh, A., 2022. Immigrant women’s perspectives on contraceptive counselling provided by midwives in Sweden – a qualitative study. Sexual and Reproductive Health Matters 30. doi: 10.1080/26410397.2022.2111796

Kopp Kallner, H., Thunell, L., Brynhildsen, J., Lindeberg, M., Gemzell Danielsson, K., 2015. Use of contraception and attitudes towards contraceptive use in Swedish women - a nationwide survey. PLOS ONE 10:e0125990. doi: 10.1371/journal.pone.0125990

Levesque, J.-F., Harris, M.F., Russell, G., 2013. Patient-centred access to health care: conceptualising access at the interface of health systems and populations. International Journal for Equity in Health 12:18. doi: 10.1186/1475-9276-12-18

Newton-Levinson, A., Griffin, K., Blake, S.C., Swartzendruber, A., Kramer, M., Sales, J.M., 2024. “I probably have access, but I can’t afford it”: expanding definitions of affordability in access to contraceptive services among people with low income in Georgia, USA. BMC Health Services Research 24. doi: 10.1186/s12913-024-11133-6

Reed, R., Osby, O., Nelums, M., Welchlin, C., Konate, R., Holt, K., 2022. Contraceptive care experiences and preferences among Black women in Mississippi: a qualitative study. Contraception 114:18–25. doi: 10.1016/j.contraception.2022.05.009

Ross, L.J., Solinger, R., 2017. Reproductive justice: an introduction. University of California Press, Oakland, California.

Rutgers, 2024. Anticonceptie regelen. Available from: https://seksualiteit.nl/onderwerpen/anticonceptie/anticonceptie-regelen/

Sedlecky, K., Rašević, M., Bitzer, J., 2020. Education and training of health care workers for contraceptive service delivery in 21 countries across Europe. Sexual & Reproductive Healthcare 24. doi: 10.1016/j.srhc.2020.100498

Spaich, S., Weiss, C., Sütterlin, M., 2019. Altered patient perceptions and preferences regarding male and female gynecologists: a comparison between 1997 and 2018. General Gynecology 300:1331–41. doi: 10.1007/s00404-019-05315-5

Teal, S., Edelman, A., 2021. Contraception selection, effectiveness, and adverse effects. JAMA 326:2507. doi: 10.1001/jama.2021.21392

United Nations. Report of the International Conference on Population and Development: Cairo, 5–13 September 1994. New York: 1995.

van Ditzhuijzen, J., Olofsen, S., Knibbeler, R., Vlugt, I.v.d. Het eerste anticonceptieconsult bij de huisarts: tevredenheid, verwachtingen, en ervaringen van jonge vrouwen. Utrecht: Rutgers; 2021.

Van Goethem, M., Mortelmans, D., Bruyninckx, E., Verslegers, I., Biltjes, I., Van Hove, E., De Schepper, A., 2003. Influence of the radiographer on the pain felt during mammography. European Radiology 13. doi: 10.1007/s00330-002-1686-6

Wagemakers, A., Vaandrager, L. Een nieuwe rol voor de verloskundige. Onderzoek naar kansen en belemmeringen voor de Wageningse verloskundigen. Wageningen: Wetenschapswinkel Wageningen; 2015 Contract No.: 309.

Walker, B., Wisniewski, J., Tinkler, S., Torres, J., Sharma, R., 2024. Identity and access: gender-based preferences and physician availability in primary care. Journal of Economic Behavor & Organization 224:1022–36. doi: 10.1016/j.jebo.2024.07.009

Walker, S.H., Hooks, C., Blake, D., 2021. The views of postnatal women and midwives on midwives providing contraceptive advice and methods: a mixed method concurrent study. BMC Pregnancy and Childbirth 21. doi: 10.1186/s12884-021-03895-2

Whelehan, P., Evans, A., Ozakinci, G., 2016. Client and practitioner perspectives on the screening mammography experience. European Journal of Cancer Care 26:e12580. doi: 10.1111/ecc.12580

WHO. Task sharing to improve access to family planning/contraception. Geneva, Switzerland: Department of Reproductive Health and Research, World Health Organization; 2017.

Wingo, E., Sarnaik, S., Michel, M., Hessler, D., Frederiksen, B., Kavanaugh, M.L., Dehlendorf, C., 2023. The status of person-centered contraceptive care in the United States: results from a nationally representative sample. Perspectives on Sexual and Reproductive Health 55:129–39. doi: 10.1363/psrh.12245

Zell, B., Effland, K., Snyder, M., Hays, K., Gordon, W., 2024. Prescriptive authority for direct entry midwives in Washington state: increasing client access to contraception. Journal of Midwifery & Women’s Health. doi: 10.1111/jmwh.13606

