## Appendix B for "Experience of contraceptive care by midwives for nonpostpartum individuals in the Netherlands: a mixed methods study"

**Appendix B: Interview guide contraception at the midwife**

- Thank you for wanting to participate in this interview. I emailed you the information letter before this interview. Have you had time to read it through? Do you have any questions about that?
- Have you filled out the consent form?
- Getting contraception from the midwife is fairly new in the Netherlands, which is why we are doing this study. I will first ask you about previous experience with getting contraception and in the second part we will go more into your experience at the midwife. The interview will last about 45 minutes.
- There are no right or wrong answers.
- If I ask a question that you would prefer not to answer, please let me know. And if for any reason you want to stop or pause during the interview, you are welcome to say as well.
- The interview will be anonymized. Your name will not be mentioned anywhere and all information that can be traced back to you will not be included in the typed interview.
- Do you have any questions before we start?
- Then I will start the recording and start with the questions.

### Sociodemographic characteristics

- What is your age? How old are you?
- Where were you born? And where did you grow up?
- Where do you live now?
- Are you in a relationship? (Do you live together? Are you married?)
- Have you ever been pregnant? (Do you have kids?)
- What kind of school did you go to after primary school? What school did you finish?
- Do you work? (What kind of work do you do?)
- Were you raised with a religion? (Which one?)

### Contraception life course

- I am curious about the different contraceptive methods you have used in your life and how you experienced the access to them. Let’s list all the contraception you have used, and I’ll ask questions about each. (*Draw out.)*
  - How old were you when you first started using contraception? *(including condoms)*
  - Why did you want to start?
  - What form of contraception were you using at the time?
    - Here is a list of different methods (*with Rutgers visual aid*)
  - **Where did you get [contraceptive method] back then? (e.g., general practitioner, gynaecologist, abortion clinic, midwife) *Add place/caregiver to life course.***
    - Why did you decide to go there?
    - How was your experience there?
  - **How much did it cost you to get [contraceptive method] back then?**
    - Was part of it covered by your health insurance? Did you know?
    - Did costs play a role for you at the time? If so, what kind of role?
  - Does it play a role for you if health care statements show that you use contraception *(e.g., if parents/partner pay for health insurance)?* If so, what kind of role?
  - How long did you use [contraceptive method] then? / How old were you when you stopped using it?
    - If stopped: Why did you want to stop using the contraception? Did you discuss this with the person who prescribed it?
  - Did you start using something else after that to prevent getting pregnant?
- *Repeat the above questions up to and including the current method:*
  - *1. Starting age, 2. Reason start, 3. Which contraceptive method (including condoms), 4. Place of access, why there, experience. 5. Costs. 6. Stopping age, 7. Reason stop/discussed with HCP. Also note if nothing has been used for a period.*
- Which midwife practice did you visit?

### Experience with the midwife

Now I would like to talk about your experience with contraceptive care at the midwife.

- First, I'm curious about how you chose contraception for the midwife.
  - **How did you know you could go to the midwife for contraception?**
    - Where did you find (your) information?
    - What did you think about the information about contraceptive care at the midwife?
      - To what extent was this information understandable to you? What would you change about this?
    - How did you make an appointment? What did you think of the availability?
      - How fast were you able to get an appointment?
    - What kind of appointment did you have? *Insertion and/or consultation.*
  - **What made you choose to go to the midwife for birth control?**
    - *Personal, social, cultural norms and values; Gender (caregiver)*
- **What did you expect from the contraceptive care at the midwife? *(first open, then ask further questions if necessary)***
  - How many/which different forms of contraception did you expect the midwife to inform you about?
  - What kind of information about the different methods? *(e.g., reliability, side effects, use, period, costs, STI’s, appropriate to personal (medical) situation)*
  - To what extent did you expect the midwife to be able to help you make your choice? Did you expect that your final choice would be made by you, the midwife or together?
    - Did the midwife ask you why you chose [self chosen contraception]?
  - *Expectations of satisfaction, nerves/tension, asking about sexual relationships, asking about reason for AC use, trust that midwife wouldn't tell anyone else.*
- **Then I am curious how the contraceptive care at the midwife went.**
  - How did you go there (e.g., by tram, bicycle, on foot, etc.)? Was it easy to reach?
  - Did you have a (telephone) intake before? If so, how did that go?
    - Did they ask about STI test? Pregnancy test?
  - **How did it go during your appointment? *(first open, then ask further questions if necessary)***

**Can you take me along through your appointment from the moment you arrived?**

- Waiting room: What was the waiting room like? How long did you have to wait? Was it busy/lot of other people?
- Before: where did you sit? On the bench or on a different chair?
- Insertion
- Ultrasound?
- After
- How long did the appointment take?
- How many/which different forms of contraception did the midwife give you information about?
  - What kind of information about the different methods? *(e.g., reliability, side effects, use, bleeding pattern, costs, STIs, appropriate to personal (medical) situation)*
- How did you choose the method you finally chose? (guidance from midwife, self, together)
- In addition to the chosen form of contraception, did you also receive information about the use of condoms to prevent sexually transmitted infections?
- To what extent did you understand all the information you received from the midwife? And what the midwife did?
  - How comfortable did you feel asking the midwife all your questions and raising your concerns?
- After the first meeting with the midwife, was there a follow-up appointment (physical or telephone) to discuss whether you were satisfied with the contraceptive method?
- To what extent did that match what you expected?
  - Did you feel a need for a follow up?
- **How did you experience contraceptive care at the midwife? *(first open, then ask questions)***
  - What did you appreciate about the midwife?
  - What did you dislike?
    - How could that be better?
  - To what extent did the midwife meet your needs?
    - Would you recommend the midwife for contraception to a friend?
      - Why? Why not?
      - Specifically, this one or midwives in general?
  - Would you recommend the midwife to your younger self?
  - *If not named, ask questions about professionalism of midwife, sufficient time, social skills (friendly, open, safe, respectful), continuity (follow-up)*
- Costs / covered by health insurance
  - Method
  - Insertion
  - Ultrasound
  - Follow-up
- What was it like for you to see a midwife when you are not pregnant?
- Are there any specific things that need to be changed regarding contraception at the midwife?
- **How did this experience compare to your previous experiences with getting birth control?** *Depending on previous experience:*
- General practitioner, gynaecologist, abortion doctor
  - In terms of accessibility, degree of acceptance, accessibility, affordability, and appropriateness
  - Gender of healthcare provider
- Where would you go next time? For example, for removal or getting new contraception
  - Why would you take the contraception out? *E.g., desire to have children?*
- What else do you need to get contraception and use it properly?
  - Any other wishes for contraception and/or access to contraception?

### Closing

- This was my last question. Is there anything else you would like to add?
- Do you have any questions for me?
- Then I will stop the recording now.
- I want to thank you very much for your cooperation and time. Here is the gift card.
