## Appendix A for "Experience of contraceptive care by midwives for nonpostpartum individuals in the Netherlands: a mixed methods study"

**Appendix A: Survey contraception at the midwife**

| **1** | **Consent** | |
| --- | --- | --- |
|  | We check if you can take part and we ask for your consent. | |
| 1.1 | How old are you? | [text box] years |
| 1.2 | Did you give birth in the past six months? | - Yes - No |
| 1.3 | I give the researchers permission to collect and use my data as been described in the information letter.  If you don’t want to give permission, you cannot complete the survey. | - Yes |
| **2** | **Personal characteristics** | |
|  | The next questions are about personal characteristics. | |
| 2.1 | What is the highest level of education that you have completed? | - Primary school - Practical training - Pre-vocational secondary education (VMBO) - Senior general secondary education (HAVO) - Pre-university secondary education (VWO) - Secondary vocational education (MBO) - Higher professional education (HBO) - University education (WO) - Other, please describe:... - Do not know - Rather not say |
| 2.1.1 | *[If 2.1 = Other, please describe:...]*  Other, please describe:... | [text box] |
| 2.2 | In which country were you born? | - Netherlands - A different country in Europe - Turkey - Morocco - Surinam - Dutch Caribbean (formerly Netherlands Antilles) - Indonesia - A different country outside of Europe - Do not know - Rather not say |
| 2.3 | In which country was your mother born? | - Netherlands - A different country in Europe - Turkey - Morocco - Surinam - Dutch Caribbean (formerly Netherlands Antilles) - Indonesia - A different country outside of Europe - Do not know - Rather not say |
| 2.4 | In which country was your father born? | - Netherlands - A different country in Europe - Turkey - Morocco - Surinam - Dutch Caribbean (formerly Netherlands Antilles) - Indonesia - A different country outside of Europe - Do not know - Rather not say |
| 2.5 | What is your religion? | - No religion - Buddhism - Christianity (e.g. Roman Catholic or Protestant) - Hinduism - Islam - Judaism - Other, please describe:... - Do not know - Rather not say |
| 2.5.1 | *[If 2.5 = Other, please describe:...]*  Other, please describe:... | [text box] |
| 2.6 | Which description best resembles your view about the current income of your household right now? | - Living very comfortably on present income - Living comfortably on present income - Coping on present income - Finding it difficult on present income - Finding it very difficult on present income - Do not know - Rather not say |
| 2.7 | Are you in a relationship or do you have a regular partner? | - Yes - No |
| 2.8 | Have you visited a midwife before? | - Yes - No - Do not know - Rather not say |
| **3** | **Previous experience with contraception access** | |
|  | The next questions are about your earlier experiences with accessing contraception. | |
| 3.1 | Have you received advice about contraception from a healthcare provider before? A healthcare provider is for example a doctor or a nurse. | - Yes - No, this is the first time |
| 3.1.1 | *[If 3.1 = Yes]*  From which healthcare provider did you receive advice about contraception before? (multiple answers possible) | - General practitioner (GP) - Gynaecologist - Abortion clinic - Midwife - Sense nurse or doctor - Other, please describe:... - Do not know - Rather not say |
| 3.1.1.1 | *[If 3.1.1 = Other, please describe:...]*  Other, please describe:... | [text box] |
| 3.2 | Have you used contraception before? | - Yes - No, this is the first time |
| 3.2.1 | Which healthcare provider(s) have given you contraception before? (multiple answers possible) | - General practitioner (GP) - Gynaecologist - Abortion clinic - Midwife - Sense nurse or doctor - Other, please describe:... - Do not know - Rather not say |
|  | *[If 3.2.1 = Other, please describe:...]*  Other, please describe:... | - [text box] |
| 3.2.2 | *[If 3.2 = Yes]*  Were you using contraception in the month before your appointment at the midwife? | - Yes - No |
| **4** | **Talks about contraception** | |
|  | The next questions are about who you talked with about contraception, and how important their opinion is to you. | |
| 4.1 | *[If 2.7 = Yes]*  Did you talk about contraception with your partner prior to this appointment? | - Yes - No - Do not know - Rather not say |
| 4.2 | *[If 2.7 = Yes]*  How important is your partners opinion about contraception to you? | - Very important - Important - Somewhat important - Not important - Do not know - Rather not say |
| 4.3 | Did you talk about contraception with other people prior to this appointment? | - Yes - No - Do not know - Rather not say |
| 4.3.1 | *[If 4.3 = Yes]*  With whom? (multiple answers possible) | - Family - Friends - Colleagues - Acquaintances - Classmates - Do not know - Rather not say |
| 4.4 | How important is the opinion of other people about contraception to you? | - Very important - Important - Somewhat important - Not important - Do not know - Rather not say |
| **5** | **Your appointment** | |
|  | The next questions are about the reason of your appointment, scheduling the appointment, and the type of appointment. | |
| 5.1 | Why did you have an appointment for contraception right now? (For example, because you were unhappy about the contraception you were using or because you wanted to start again) | [text box] |
| 5.2 | How easy was it to make an appointment at the midwife? | - Very easy - Easy - Moderately easy - Not easy - Do not know - Rather not say |
| 5.3 | When you made an appointment, when were you able to visit the midwife for contraception? | - The same day - Within a few days - Within a week - Within two weeks - After two weeks - Do not know - Rather not say |
| 5.4 | How easy was it for you to come to the appointment (e.g. walking, biking, public transport, car, etc.)? | - Very easy - Easy - Moderately easy - Not easy - Do not know - Rather not say |
| 5.5 | How understandable was the information about contraception at the midwife prior to your appointment? For example about different methods of contraception or the appointment itself. | - Very understandable - Understandable - Somewhat understandable - Not understandable - Do not know - Rather not say |
| 5.6 | Why did you go to the midwife? | - I only went to the midwife for advice about contraception - I only went to the midwife to get contraception (insertion or prescription) - I went to the midwife both for advice and to get contraception (insertion or prescription) |
| 5.7 | What happened during your appointment? | - I only got advice about contraception - I only got contraception (insertion or prescription) - I both got advice and I got contraception (insertion or prescription) |
| 5.8 | How did you know that you could go to the midwife for contraception? (multiple answers possible) | - Through family, friends, acquaintances - Through social media - Through the midwife - Through the general practitioner (GP) - Other, please describe:... - Do not know - Rather not say |
| 5.8.1 | *[If 5.8 = Other, please describe:...]*  Other, please describe:… | [text box] |
| 5.9 | At which midwife practice did you have an appointment? | - Wereldkind, Alphen aan den Rijn - Echo, Amsterdam - Femme, Amsterdam - Nova, Amsterdam - Vida, Amsterdam - Geboortehuis Bussum - Carmenta, Den Haag - Ella, Den Haag - Femme, Den Haag - Geboortecentrum Life, Den Haag - Anticonceptiespecialist Wendy, Groningen - De Bakermat, Wageningen - Wel en Wee, Winterswijk - De Kiem, Zwolle - Other, please describe:... - Rather not say |
|  | *[If 5.9 = Other, please describe:...]*  Other, please describe:… | [text box] |
| 5.10 | How long ago was your appointment for getting advice on or receiving contraception? | - Less than 1 week ago - Less than 2 weeks ago - Less than 1 month ago - Less than 2 months ago - More than 2 months ago - Do not know - Rather not say |
| 5.10.1 | How long ago was your appointment? | [text box] |
|  | The next questions are about your sex preference for a health care provider in contraceptive care. | |
| 5.11 | Was the midwife you had an appointment with a woman or a man? | - Woman - Man |
| 5.12 | Who do you prefer to go to for advice about contraception? | - A woman - A man - I do not have a preference - Do not know - Rather not say |
| 5.12.1 | *[If 5.12 = A woman]*  How important is it for you that the healthcare provider is a woman for advice about contraception? | - Very important - Important - Somewhat important - Not important - Do not know - Rather not say |
| 5.12.2 | *[If 5.12 = A man]*  How important is it for you that the healthcare provider is a man for advice about contraception? | - Very important - Important - Somewhat important - Not important - Do not know - Rather not say |
| 5.13 | Who do you prefer to go to for getting contraception? For example for insertion or prescription. | - A woman - A man - I do not have a preference - Do not know - Rather not say |
| 5.13.1 | *[If 5.13 = A woman]*  How important is it for you that the healthcare provider is a woman for the insertion or prescription of contraception? | - Very important - Important - Somewhat important - Not important - Do not know - Rather not say |
| 5.13.2 | *[If 5.13 = A man]*  How important is it for you that the healthcare provider is a man for the insertion or prescription of contraception? | - Very important - Important - Somewhat important - Not important - Do not know - Rather not say |
| **6** | **Expectations and experiences** | |
|  | The next questions are about different methods of contraception. | |
| 6.1 | Before going to the midwife, which contraception did you prefer? (max. 3 answers) | - I did not have a preference yet - Combined pill - Progesteron-only pill - Intrauterine system (IUS) (hormonal) - Intrauterine device (IUD) (copper) - Contraceptive implant - Contraceptive patch - Vaginal ring - Contraceptive injection - Male condom - Female condom - Diaphragm - Natural family planning: for example tracking the ovulation with calendar or a fertility app - Withdrawal method (pulling out) - Sterilisation - Do not know - Rather not say |
| 6.2 | Which forms/methods of contraception did you expect the midwife to tell you about? (multiple answers possible) | - Combined pill - Progesteron-only pill - Intrauterine system (IUS) (hormonal) - Intrauterine device (IUD) (copper) - Contraceptive implant - Contraceptive patch - Vaginal ring - Contraceptive injection - Male condom - Female condom - Diaphragm - Natural family planning: for example tracking the ovulation with calendar or a fertility app - Withdrawal method (pulling out) - Sterilisation - Do not know - Rather not say |
| 6.3 | Which forms/methods of contraception did the midwife tell you about? (multiple answers possible) | - Combined pill - Progesteron-only pill - Intrauterine system (IUS) (hormonal) - Intrauterine device (IUD) (copper) - Contraceptive implant - Contraceptive patch - Vaginal ring - Contraceptive injection - Male condom - Female condom - Diaphragm - Natural family planning: for example tracking the ovulation with calendar or a fertility app - Withdrawal method (pulling out) - Sterilisation - Do not know - Rather not say |
| 6.4 | Which form of contraception did you end up choosing? (max 2 answers) | - Combined pill - Progesteron-only pill - Intrauterine system (IUS) (hormonal) - Intrauterine device (IUD) (copper) - Contraceptive implant - Contraceptive patch - Vaginal ring - Contraceptive injection - Male condom - Female condom - Diaphragm - Natural family planning: for example tracking the ovulation with calendar or a fertility app - Withdrawal method (pulling out) - Sterilisation - Do not know - Rather not say |
| 6.5 | Was the contraceptive method you chose the same as the method you thought of prior to your appointment? | - Yes - No - I did not know which contraception I wanted before the appointment - Do not know - Rather not say |
|  | The next questions are about subjects the midwife told and asked about. | |
| 6.6 | *[If 5.7 = I only got advice about contraception. i.e., appointment was advice only]*  What did you expect the midwife to tell you about different methods of contraception? (multiple answers possible)  *[If 5.7 =! I only got advice about contraception. i.e., appointment included insertion/prescription]*  What did you expect the midwife to tell you about the contraception you chose? (multiple answers possible) | - What the chances are of becoming pregnant - How it works - How to use it - How it might change my period - Side effects - Costs - If you can get an STI while using it - If it fits my personal situation/body - What if something goes wrong - Do not know - Rather not say |
| 6.7 | What did the midwife tell you about? (multiple answers possible) | - What the chances are of becoming pregnant - How it works - How to use it - How it might change my period - Side effects - Costs - If you can get an STI while using it - If it fits my personal situation/body - What if something goes wrong - Do not know - Rather not say |
| 6.8 | Did you receive information about the use of condoms to prevent STI? | - Yes - No - Do not know - Rather not say |
| 6.9 | How understandable was the information of the midwife during your appointment? | - Very understandable - Understandable - Somewhat understandable - Not understandable - Do not know - Rather not say |
| 6.10 | Did the midwife ask about your (sexual) relationships? | - Yes - No - Do not know - Rather not say |
| 6.11 | Did you expect the midwife to ask about your (sexual) relationship(s)? | - Yes - No - Do not know - Rather not say |
| 6.12 | Did the midwife ask you about the reason for your appointment? | - Yes - No - Do not know - Rather not say |
| 6.13 | Did you expect the midwife to ask about the reason for your appointment? | - Yes - No - Do not know - Rather not say |
|  | The next questions are about how you chose a contraception method. | |
| 6.14 | Did you expect the midwife to give an opinion about which contraception is best for you? | - I very much expected that the midwife would give their opinion - I somewhat expected that the midwife would give their opinion - I did not really expect that the midwife would give their opinion - I did not expect the midwife that the midwife would give their opinion - Do not know - Rather not say |
| 6.15 | To what extent did the midwife give an opinion about which contraception is best for you? | - The midwife very strongly gave their opinion - The midwife somewhat gave their opinion - The midwife did not really give their opinion - The midwife did not give their opinion - Do not know - Rather not say |
| 6.16 | Did you expect to be able to make the choice for contraception by yourself (without the opinion of the midwife)? | - I expected to be able to make the choice myself - I somewhat expected to be able to make the choice myself - I did not really expect to be able to make the choice myself - I did not expect to be able to make the choice myself - Do not know - Rather not say |
| 6.17 | Did you make the choice for the type of contraception yourself? | - I completely made the choice myself - I almost completely made the choice myself - I mostly did not make the choice myself - I did not make the choice myself - Do not know - Rather not say |
| 6.18 | Did the appointment go like you expected it to go? | - Yes - A little bit - No - Do not know - Rather not say |
|  | The next question are about a follow-up appointment. | |
| 6.19 | Did you want a follow up appointment (in person or by phone) where you could discuss how things are going with the contraception and/or to ask more questions? | - Yes - A little bit - No - Do not know - Rather not say |
| 6.20 | Was there a follow up appointment with the midwife to discuss if you were satisfied with your contraception. This could be an in-person appointment with the midwife or over the phone. | - Yes - No, I was asked, but I did not need it - No, I did not get asked - Do not know - Rather not say |
| **7** | **How was it for you?** | |
|  | The next questions are about how you felt during the appointment. | |
| 7.1 | How pleased were you about the appointment for contraception with the midwife? | - Very pleased - Pleased - Moderately pleased - Not pleased - Do not know - Rather not say |
| 7.2 | Did you feel at ease during the appointment? | - Yes - A little bit - No - Do not know - Rather not say |
| 7.3 | Did you feel like you were taken seriously by the midwife during the appointment? | - Yes - A little bit - No - Do not know - Rather not say |
| 7.4 | If you were concerned about something, did you feel like you could tell the midwife? | - Yes, I was able to express concern - No, I was not able to express concern - I did not have any concerns - Do not know - Rather not say |
| 7.5 | Were you able to ask the midwife all your questions? | - I was able to ask all questions - I was able to ask nearly all questions - I was barely able to ask any questions - I was not able to ask any questions - Do not know - Rather not say |
| 7.6 | Did you think the midwife had enough time for the appointment? | - Yes, the midwife had enough time - No, the midwife had too little time - Do not know - Rather not say |
| 7.7 | Did you trust that the midwife would not share anything about your appointment with anyone else? For example your partner, parents or other healthcare provider. | - I had lots of trust - I had trust - I had little trust - I had no trust - Do not know - Rather not say |
| **8** | **Costs** | |
|  | The next questions are about costs. | |
| 8.1 | *[If 5.7 =! I only got contraception (insertion or prescription). i.e., appointment included advice]*  Did you have to pay for the advice about contraception with the midwife? | - Yes, I had to pay everything myself - Yes, I had to pay part of it myself - No, it was fully covered by my health insurance - No, I did not have to pay anything - Do not know - Rather not say |
| 8.2 | *[If 5.7 =! I only got advice about contraception. i.e., appointment included insertion/prescription]*  Did you have to pay for your contraception method? This is for example the IUD, hormonal implant, or birth control pill itself. It is not about the prescription or insertion. | - Yes, I had to pay everything myself - Yes, I had to pay part of it myself - No, it was fully covered by my health insurance - No, I did not have to pay anything - Do not know - Rather not say |
| 8.3 | *[If 5.7 =! I only got advice about contraception. i.e., appointment included insertion/prescription]*  Did you have to pay for getting the contraception at the midwife? This is for example the insertion of an IUD or implant, or a prescription. | - Yes, I had to pay everything myself - Yes, I had to pay part of it myself - No, it was fully covered by my health insurance - No, I did not have to pay anything - Do not know - Rather not say |
| 8.4 | Does money play a role in choosing the health care provider for contraception? | - Plays a large role - Plays a small role - Plays no role - Do not know - Rather not say |
| **9** | **Final questions** | |
|  | The next questions are about what you appreciated and what can be improved. | |
| 9.1 | *[If 5.7 =! I only got contraception (insertion or prescription). i.e., appointment included advice]*  If you had to rate the advice about contraception at the midwife, which grade would you give? | - 0 = very bad - 1 - 2 - 3 - 4 - 5 - 6 - 7 - 8 - 9 - 10 = very good - Do not know - Rather not say |
| 9.2 | *[If 5.7 =! I only got advice about contraception. i.e., appointment included insertion/prescription]*  If you had to rate the insertion or prescription of contraception at the midwife, which grade would you give? | - 0 = very bad - 1 - 2 - 3 - 4 - 5 - 6 - 7 - 8 - 9 - 10 = very good - Do not know - Rather not say |
| 9.3 | What did you appreciate, before, during or after your appointment for contraception at the midwife? | [text box] |
| 9.4 | What did you not appreciate as much, before, during or after your appointment for contraception at the midwife? | [text box] |
| 9.5 | What could have been done better before, during or after your appointment for contraception at the midwife? (multiple answers possible) | - Nothing, it was a very good appointment - There could have been more attention for my personal situation - There could have been more time - There could have been more explanation about the different methods - There could have been more explanation about the reliability - There could have been more explanation about the side effects - There could have been more information about natural methods/non hormonal methods - There could have been more explanation about what to do when something goes wrong - The midwife had too much of an opinion - They did not listen to me well enough - Other, please describe:... - Do not know - Rather not say |
| 9.5.1 | *[If 9.5 = Other, please describe:...]*  Other, please describe:... | [text box] |
| 9.6 | Do you have any tips for midwifes to improve their contraceptive care? | [text box] |
| 9.7 | *[If 5.7 =! I only got contraception (insertion or prescription). i.e., appointment included advice]*  Would you recommend the midwife to a friend for advice about contraception? | - Yes - No - Do not know - Rather not say |
| 9.8 | *[If 5.7 =! I only got advice about contraception. i.e., appointment included insertion/prescription]*  Would you recommend the midwife to a friend for insertion or prescription of contraception? | - Yes - No - Do not know - Rather not say |
| 9.9 | *[If 5.7 =! I only got contraception (insertion or prescription). i.e., appointment included advice]*  If you need contraception again another time, to which healthcare provider would you go for advice? | - General practitioner (GP) - Gynaecologist - Abortion clinic - Midwife - Sense nurse or doctor - Other, please describe:... - Do not know - Rather not say |
| 9.9.1 | *[If 9.9 = Other, please describe:...]*  Other, please describe:... | [text box] |
| 9.10 | *[If 5.7 =! I only got advice about contraception. i.e., appointment included insertion/prescription]*  If you need contraception again another time, to which healthcare professional/provider would you go for retrieval/insertion/to get it? | - General practitioner (GP) - Gynaecologist - Abortion clinic - Midwife - Sense nurse or doctor - Other, please describe:... - Do not know - Rather not say |
|  | *[If 9.10 = Other, please describe:...]*  Other, please describe:... | [text box] |
| 9.11 | Are there any other things that you would like to share about your experience with contraception at the midwife? | [text box] |
|  | The next question is about abortion care at the midwife. | |
| 9.12 | Imagine you have an unwanted pregnancy that you wish to terminate. Would you want to access abortion care at your midwife? Midwifes are available by phone 24/7 for questions and they can easily come to you if you need additional support. | - Yes - No - Do not know - Rather not say |
| 9.12.1 | *[If 9.12 = Yes]*  For what reasons would you want this? | [text box] |
| 9.12.2 | *[If 9.12 = No]*  For what reasons would you not want this? | [text box] |
|  | That was the final question, thanks for taking part! On the next page, you can find instructions for requesting the VVV voucher. | |
